# Efficacy assessment of newly-designed and locally-produced filtering facemasks during the SARS-CoV-2 pandemic

**DOI:** 10.1101/2020.09.04.20188185

**Authors:** Bob Boogaard, Ali Tas, Joep Nijssen, Freek Broeren, John van der Dobbelsteen, Vincent Verhoeven, Jip Pluim, Sing Dekker, Eric Snijder, Martijn van Hemert, Sander Herfst

## Abstract

The SARS-CoV-2 pandemic resulted in shortages of production and test capacity of FFP2-respirators. Such facemasks are required to be worn by healthcare professionals when performing aerosol-generating procedures on COVID-19 patients. In response to the high demand and short supply, we designed three models of facemasks that are suitable for local production. As these facemasks should meet the requirements of an FFP2-certified facemask, the newly-designed facemasks were tested on the filtration efficiency of the filter material, inward leakage, and breathing resistance with custom-made experimental setups. In these tests, the locally-produced facemasks were benchmarked against a commercial FFP2 facemask. Furthermore, the protective capacity of the facemasks was tested for the first time with coronavirus-loaded aerosols under physiologically relevant conditions. This multidisciplinary effort resulted in the design and production of facemasks that meet the FFP2 requirements, and which can be mass-produced at local production facilities.

## INTRODUCTION

The severe acute respiratory syndrome coronavirus-2 (SARS-CoV-2) is a novel coronavirus that was first identified in December 2019 in Wuhan, China, in patients suffering from acute respiratory syndrome (2019 coronavirus disease or COVID-19) (Zhu et al., 2020). SARS-CoV-2 was declared pandemic by WHO on March 11, 2020.

Healthcare professionals who are involved in aerosol-generating procedures on COVID-19 patients are required to use FFP2-classified filter facepiece respirators for respiratory protection against SARS-CoV-2 infection. FFP2-respirators (from now on called ‘facemask’) filter at least 94% of the submicron-sized test aerosols with NaCl, according to the NEN-EN 149:2001 + A1:2009 standards (**Table 1**). These standards are used by the accredited test laboratories (the so-called notified bodies) within the European Union member states to test and certify the facemasks upon approval. According to these standards, FFP2-classified facemasks are also required to meet the criteria for inward leakage, maximal CO_2_-content of inhaled air, and breathing resistance, which are summarized in **Table 1**.

**Table 1:** Requirements for FFP2-classified FFRs according to the NEN-EN149 standards

| **Filter**<br>**penetration test** | **FIT-test** | **CO<sub>2</sub> content test** | **Breathing resistance test** |  |
| --- | --- | --- | --- | --- |
| Max. penetration<br>of 120 mg NaCl<br>test aerosol at 95<br>L min<sup>-1</sup> | Max. inward<br>leakage | Max. CO<sub>2</sub> content<br>of inhaled air | Inhalation | Exhalation |
| 6% | 8<sup>a</sup> or 11<sup>b</sup> % | <1% on average | 0.7 mbar at<br>30 L min<sup>-1</sup><br>2.4 mbar at<br>95 L min<sup>-1</sup> | 3.0 mbar at<br>160 L min<sup>-1</sup> |

<sup>a</sup> for at least 8/10 individual wearer arithmetic means
<sup>b</sup> for at least 46/50 individual exercise results (e.g. 10 subjects x 5 exercises)

As the global demand for FFP2-facemasks largely exceeded the production, distribution and test capacities of the conventional suppliers and notified bodies during the COVID-19 pandemic, we formed a Dutch collaborative initiative, consisting of the Reinier de Graaf hospital, Royal DSM, Delft University of Technology, Leiden University Medical Center and Erasmus University Medical Center, with the aim to design and produce facemasks that meet the FFP2-specifications. Those locally-produced facemasks were initially tested with custom-made developed test equipment for a NaCl penetration test, fit test, and breathing resistance test under conditions that approximate the NEN-EN149 standards before being tested and approved by a certified test laboratory. Those setups have been described in detail in Blad et al., 2020 (submitted). The facemasks were also tested on virus filtration efficiency (VFE) with the mouse hepatitis virus (MHV), a beta coronavirus that causes lethal hepatitis in mice (Gledhill et al., 1955), but that is non-pathogenic to humans. This test provides additional and biologically more relevant evidence for the filtration efficiency of submicron-sized particles by the facemask’s filter material. These studies identified mask designs that protect against coronavirus-loaded aerosols with an efficiency similar to that of a commercial FFP2-facemask.

## METHODS

### Facemask designs

Three different types of in-house designed facemasks were tested in triplicate in various custom-made experimental setups. The “Reinier 0.1” facemask (**Fig. 1a**) was constructed with a dental facemask, that consisted of two layers polypropylene nonwoven fabric (40 and 20 gr m^−2^) and a single layer of 20 g m^−2^ melt-blown fabric, to which two extra layers of spun-bond polypropylene filters (100 and 20 g m^−2^) was added. The “DSM 1.0” facemask (**Fig. 1b**) consisted of five polypropylene nonwoven filter layers that consisted of 55 g m^−2^ spunmelt, 20 g m^−2^ melt-blown, 30 g m^−2^ melt-blown, 20 g m^−2^ melt-blown, and 47 g m^−2^ spunmelt polypropylene filters. The name of this mask refers to the collaboration with the Dutch nutrition and health company Royal DSM during the development of this facemask. The “Reinier 1.0” facemask (**Fig. 1c**) consisted of three layers: two layers of spun-bond polypropylene filters (100 and 20 gr m^−2^) and a single 20 gr m^−2^ melt-blown polypropylene layer. The facemasks were benchmarked against the FFP2-certified 3M Aura 1862(+) facemask.

**Fig 1.**
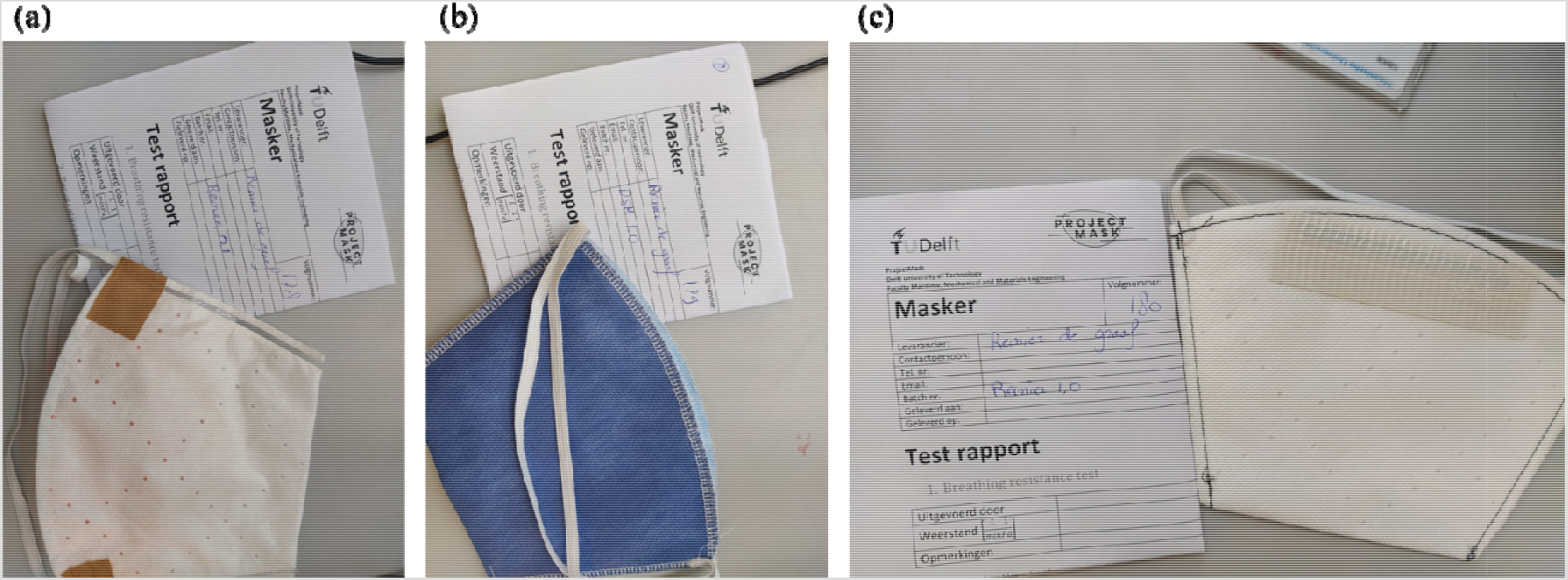
The designs of the three locally-produced facemasks with Reinier-0.1 (a), DSM-1.0 (b), and Reinier-1.0 (c).

### The dry particle penetration test

The dry environmental particle penetration test was performed to provide an initial indication of the filtering capacity of the in-house designed facemasks. The experimental setup consisted of a Solair 3200 particle counter (Lighthouse) that was connected with a particle chamber, on which a facemask was fixed in an airtight manner. The particle counter generated a flow of 56.6 L min^−1^, which created an air velocity of 0.25 m sec^−1^ through the facemask’s filter material. Particles in the range of 0.3 – 0.5μm, 0.5 – 5.0 μm, and 5.0 – 25.0 μm in size were counted. First, a reference count of the number of environmental particles (# particles_Ref_) was performed in absence of a facemask and subsequently, the number of environmental particles was counted after fixing a facemask on the particle chamber (# particles_mask_). For each of the three particle size ranges, the filter capacity was calculated according to the following formula:

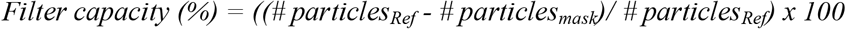

### NaCl particle penetration test

For the NaCl particle penetration test a PVC-tube system was constructed with a 90º bend, going from a vertical to a horizontal direction (the design and instructions to build are online available at https://projectmask.nl/testing/filter-material-penetration/build/). The vertical part of the tube system was connected with an Atomizer Aerosol Generator ATM 226, and the distal end of the horizontal part contained a PMMA tube with a sample holder in which a facemask was fixed in an airtight manner (**Fig. S1**). The aerosol generator produced NaCl particles from a 2% NaCl solution. The number of NaCl particles that passed through the facemask’s filter material was counted by a TSI PortaCount Pro 8030 particle counter, which generated an air velocity of 0.1 m sec^−1^ through the filter material of the facemasks. The number of particles that passed through the filter material was analysed with TSI FitPro^+^ software.

### The fittest

The fittest was performed to determine the leakage of particles around the edges of the facemasks. First, a probe was placed in the facemask by which it was connected to the PortaCount particle counter. To obtain representative results, each facemask was tested by three different individuals, who performed eight different exercises: normal breathing, deep breathing, moving the head side to side, moving the head up and down, talking, grimace, bending over and normal breathing, as described in the NEN-149 standards. The particle leakage was determined by comparing the number of particles in front and behind the facemask’s filter material and was recorded with a PortaCount particle counter. The data were analysed with the TSI FitPro^+^ software.

*The breathing resistance test*. The breathing resistance of the facemasks was determined with a custom-made experimental setup (**Fig. S2**; the design and instructions to build are online available at https://projectmask.nl/testing/breathing-resistance/build/). The facemask was placed on a manikin head (described in Zhuang and Bradtmiller et al., 2005) that was connected to a tube system, in which the pressure drop after the filter material was recorded. Before testing the facemasks, the pressure sensors were calibrated without any specimen at inlet or exit (open flow), blockage of the exit (blocked flow), or with a known flow resistance. When adding the masks, the increasing airspeed led to an increased airflow through the facemask and a pressure drop in the system, which was related to the resistance.

### Virus filtration efficiency test

As no standardized test procedures exist to determine the virus filtration efficiency of facemasks, an in-house designed experimental setup was used that consisted of a curved tube with a 0.45 m vertical and a 0.9 m horizontal part with at the distal end a sample holder for airtight placement of a facemask (**Fig. S3**.). This tube was connected to a mixing chamber, in which the virus-loaded aerosols were mixed with mist droplets, that was generated by a ultrasonic mist maker, for more efficient virus collection. The mixing chamber was connected to three SKC BioSampler impingers, in which the collected virus was impinged into 45 ml virus transport medium (VTM) (HMEM (Lonza), 12% v/v glycerol, 0.5% w/v Lactalbumin enzymatic hydrolysate, 0.02 mg ml^−1^ Polymyxin B sulfate, 0.01 mg ml^−1^ Nystatin, Penicillin/Streptomycin mixture 240/240 U ml^−1^ and 0.3 mg ml^−1^). The maximal airflow in each SKC BioSampler was 12.5 L min^−1^, generating a total maximal flow of 37.5 L min^−1^. The facemasks were challenged with mouse hepatitis virus (MHV), a beta coronavirus that causes hepatitis in mice, to be able to perform these tests under laboratory biosafety level (BSL) 2 conditions. 10^8^ plaque-forming units (PFUs) of MHV were aerosolized with an Aerogen Solo nebulizer, originally used to nebulize medication in hospital-settings, in the vertical part of the tube, directed through the bend, and subsequently the horizontal part of the tube system with the airtight fixed facemask. The facemasks were placed in the sample holder in an airtight manner between two sanitary rings with a diameter of 40 mm, which created an air velocity of 0.42 m sec^−1^ at a continuous airflow of 31.5 L min^−1^ (3 × 12.5 L min^−1^ minus 6 L min^−1^ for the mist maker). The self-made facemasks were tested in triplicate and benchmarked against the FFP2-certified 3M Aura 1862(+) facemask. The aerosolized virus was also collected in the absence of a facemask as a reference. Between each test, the tube system was flushed with HEPA-filtered air for 15 min at a flow of 37.5 L min^−1^.

*Virus quantification*. Viral RNA copy number was quantified by a quantitative reverse-transcription polymerase chain reaction (RT-qPCR) analyses and the number of infectious virus particles by plaque assays.

For RT-qPCR, viral RNA was isolated from 135 μl VTM with the QIAamp Viral RNA Mini Kit (Qiagen) according to the manufacturer’s protocol. Equine arteritis virus (EAV) was added to the lysis buffer as an internal control for the RNA isolation and RT-qPCR efficiency as described in Scheltinga *et al*., 2005. The isolated RNA was converted to copy DNA (cDNA) and quantified in a TaqMan Fast Virus 1-step master mix (Applied Biosystems) in the presence of 450 nM primers (MHV-FPr1: ACGCCGCCTTATTAAAGATG, MHV-RPr1: GGCATAGCACGATCACATTT) and 200 nM probe (TexRed-TCCTGTACTCATGGGTT GGGACTATCC-BHQ2) that targets the viral gene *nsp12*, coding for RNA dependent RNA polymerase (RdRp). The primers and probe for the EAV internal control were also added and have been described in Loens *et al*., 2012. The reactions were run in a CFX384 Q-PCR Thermocycler (BioRad) with a two-step protocol: Cycle 1 (1x): 50 ºC, 5 min. and 95 ºC for 20 sec., and Cycle 2 (45x): 95 ºC for 5 sec. and 60 ºC for 30 sec. An MHV standard was generated by *in vitro* transcription with the T7 mMessage mMachine kit (ThermoFisher) according to the manufacturer’s protocol and was also analysed as described above to determine the copy number of viral RNA in the samples.

The number of infectious virus particles was determined by a plaque assay. 8 × 10^5^ 17CL1 cells, derived from mouse (*Mus musculus*) BALB/c fibroblasts, were seeded in six-wells plates and incubated overnight at 37ºC. The cells were inoculated by incubation with an undiluted or one of the diluted MHV samples from a ten-fold serial dilution for 1 hr at 37ºC. After inoculation, the cells were washed twice with PBS, overlaid with an Avicell (Sigma) overlay, and incubated for an additional 24 hrs at 37ºC. The cells were subsequently fixated with 3.4% formaldehyde in PBS for 1 hr at room temperature and stained with 0.75% crystal violet staining solution for 5 min at room temperature. After removal of the staining solution, the wells were washed with water, and the number of plaques was counted.

## RESULTS AND DISCUSSION

### NaCl particles were more efficiently filtered than the environmental particles

To obtain a first indication of the filtering capacities of the three types of locally-produced facemasks, the filtration of dry environmental particles in a size range of 0.3 – 25 μm was determined using the Solair 3200 particle counter with and without airtight placement of one of the facemasks on the particle chamber. When the FFP2-certified facemask was placed on the particle chamber, 99.4±0.08% of the environmental particles of between 0.3-0.5 μm were filtered, and 98.9±0.27% and 95.8±1.55% of the particles of respectively 0.5-5.0 and 5.0-25.0 μm (**Fig. 2**). Varying results were obtained when the newly designed facemasks were tested in this setup. The Reinier 0.1-facemasks filtered 84±0.34% of the 0.3–0.5 μm particles, and 94.4±0.74% and 97.9±1.66% of 0.5–5.0 and 5.0–25.0 μm particles respectively. The DSM 1.0-facemasks showed stable filtering performances and filtered around 98% of the environmental particles in the three size ranges. The Reinier 1.0-facemasks on the other hand showed variable filtering performances. 0.3-0.5 μm and 0.5-5.0 μm particles were filtered for 91.4±0.25% and 96.6±0.88% respectively, while particles of the 5.0-25.0 μm size range were filtered for 94.5±7.90%. The reason for the varying filtering performances in the last particle size range is unclear as the particles smaller than 5.0 μm were filtered to a similar extent throughout the experiment. This dataset indicates that the Reinier 0.1-facemask was least capable of filtering particles of between 0.3–0.5 μm, and that the DSM 1.0-facemask performed most similar to the FFP2-certified mask.

The particle filtration efficiencies of the three facemask designs were further tested under conditions that closely approximate the NEN-149 standards with aerosolized NaCl particles. According to these standards, FFP2-facemasks are required to filter at least 94% of the NaCl particles. In our experimental setup, the FFP2-certified facemask filtered 98.13±0.84% of the NaCl particles (**Fig 3**). The Reinier-0.1 and 1.0 facemasks filtered respectively 98.9±0.42% and 99.3±0.36% of the NaCl particles. This is significantly more than observed for environmental dry particles between 0.3 – 5.0 μm, which was even below the FFP2 filtration threshold of 94% (**Fig. 2**). The Reinier-0.1 and −1.0 facemasks contained a single melt-blown polypropylene layer and the above-described observations indicate that the two layers nonwoven fabric of the three-layer dental facemask of the Reinier-0.1 model could be removed without affecting the facemask’s filtering performance. The DSM 1.0-facemasks, the only type with three layers of melt-blown polypropylene filters, showed the highest NaCl-particle filtration efficiency of 99.83±0.12% (**Fig. 3**). This model also performed best in the environmental particle filtration test (**Fig 2**).

**Fig 2.**
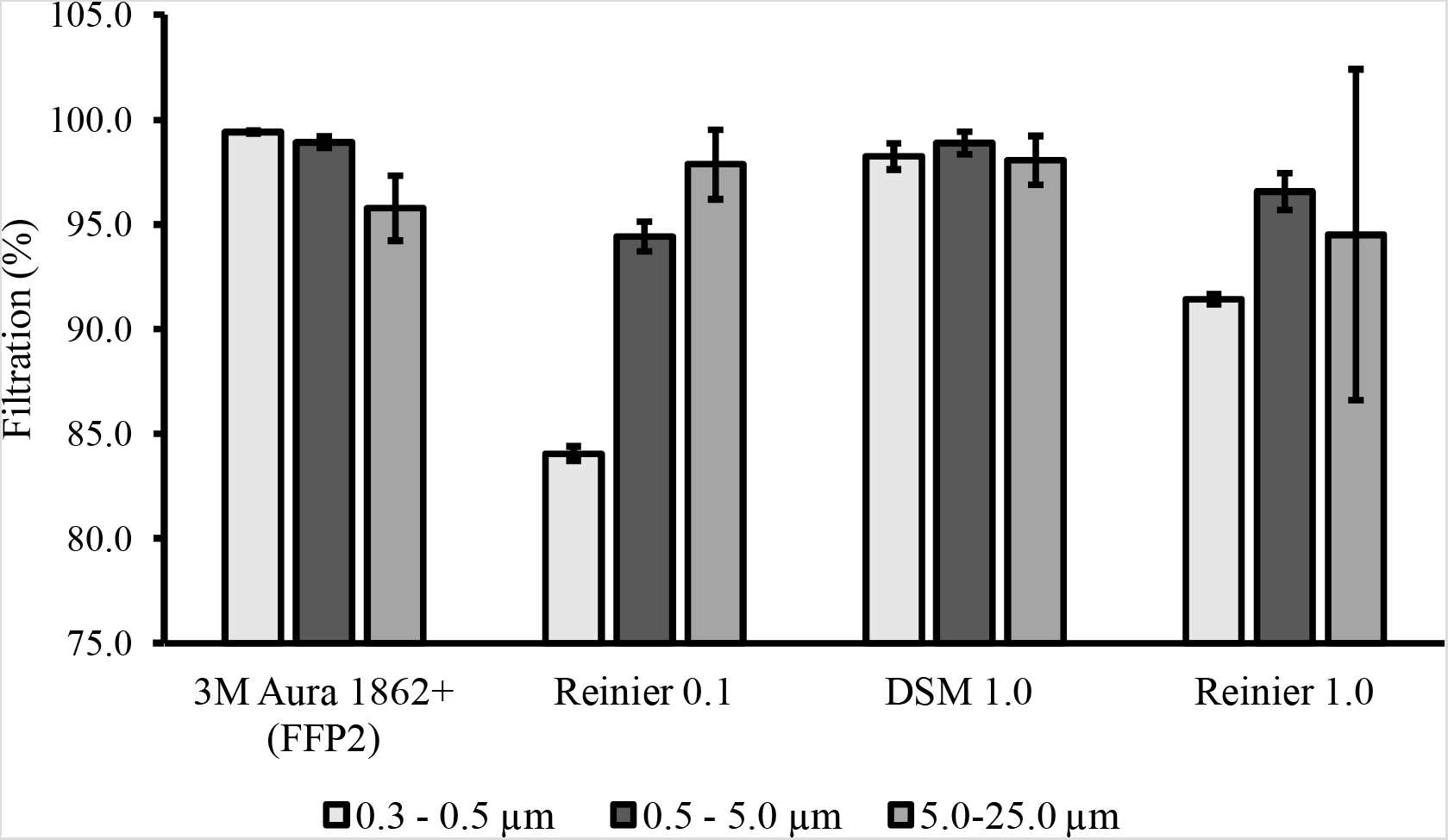
Filtration efficiency of environmental dry particles by the locally-produced facemasks and benchmarked against an FFP2-certified facemask.

**Fig 3.**
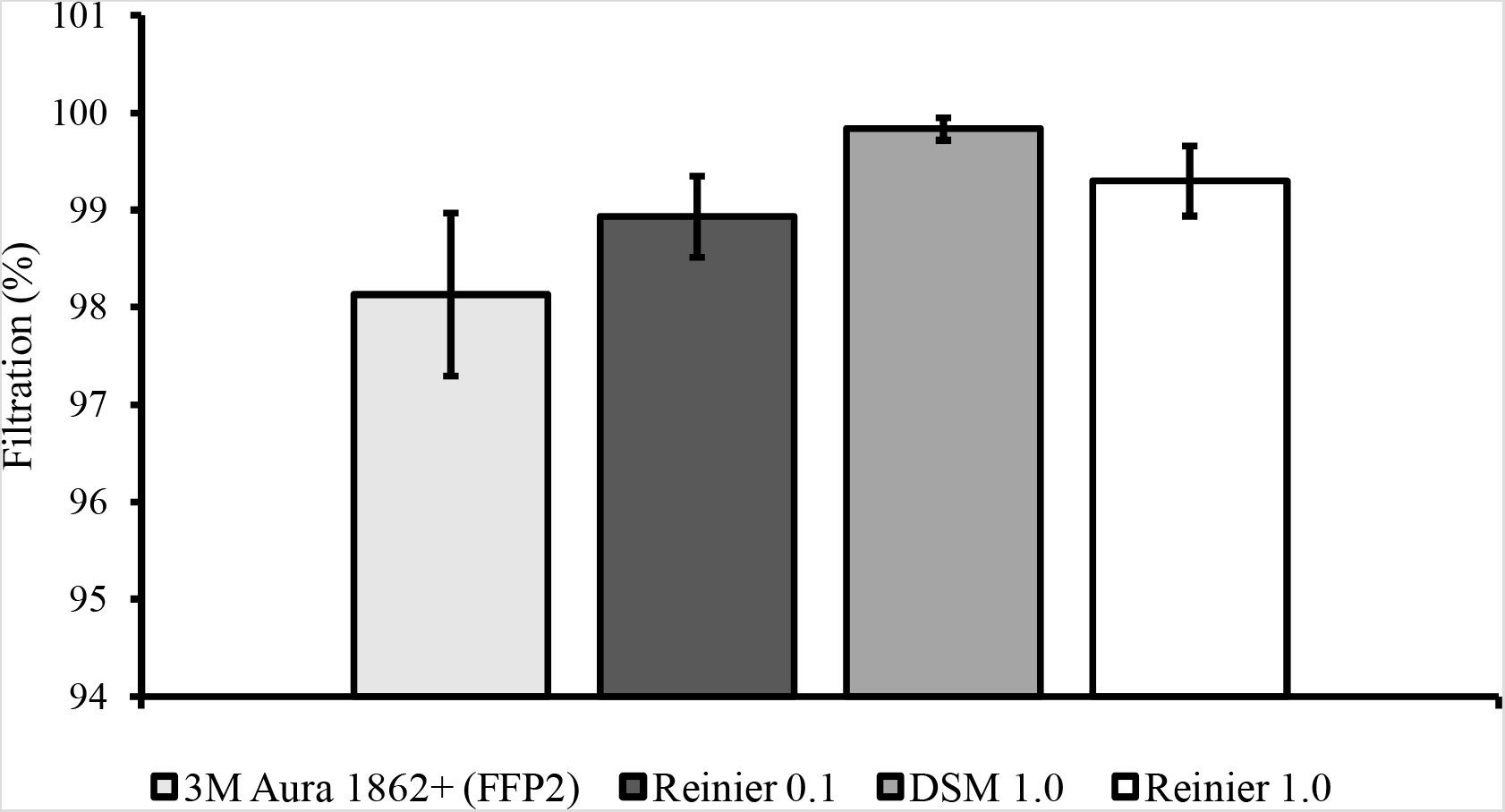
Filtration of NaCl-particles by the self-designed facemasks close to the NEN-149 standard conditions.

The observed discrepancy between the two filtration tests could be explained by varying volumetric flows and air velocities between the different experiments, as it has also been shown by others that these factors affect the facemask’s filtering performances (Rengasamy et al., 2010, 2011, Gao et al., 2016, Mukhametzanov et al., 2016). The air velocity during the environmental particle penetration test was 2.5 times higher than during the filtration test of NaCl particles and stresses the importance of standardized conditions when assessing the filter capacity of facemasks. Currently, the filtration of NaCl particles is the golden standard when testing the facemask’s filter capacity for its certification and is considered in the literature as most conservative (Rengasamy et al., 2017). The NaCl-particle filtration efficiency of the Reinier 0.1- and DSM 1.0-facemasks was also verified by notified bodies and showed similar filtration efficiencies as in this study (**Fig. S4 and S5**.), confirming that the custom-made setup indeed results in similar measurements as under the conditions of the NEN149 standards. Hence, a good indication has been obtained that the filter capacity of the three types of locally-produced facemasks meets the FFP2 requirement under standardized conditions.

### The prototype facepiece facemasks filter at least 98% of virus-loaded aerosols

To provide biologically-relevant measurements regarding their protective ability against virus-loaded aerosols, the facemasks were challenged with MHV, a beta coronavirus that infects mice. 10^8^ PFU of MHV was aerosolized, passed through a tube with and without airtight fixation of a facemask after which the air, that went through the filter material was sampled and analysed for the presence of the virus. The high number of PFUs was required to observe at least 2 logs decrease in the number of PFUs, equivalent to 99% filtration, as it was expected that most virus particles might be lost after nebulization into the tube system. The three types of facemask were tested in two separate sessions, which resulted in two independent datasets (**Fig. 4 and 5**) In the no mask-control, MHV was aerosolized into the tube system and sampled in absence of a facemask in the sample holder to determine the maximum amount of virus that could be recovered from the air. This resulted in virus recovery of 4.09 ± 0.21 log PFU ml^−1^ in the first - and 5.89 ± 0.21 log PFU ml^−1^ in the second session (**Fig. 4a and 5a**), indicating that the virus collection efficiency in the latter session was higher than in the first session. Nevertheless, the collection efficiency of infectious virus particles was high enough to be able to observe at least a 2-logs decrease in virus collection after placing a facemask in the sample holder. When an FFP2-certified facemask was placed in the sample holder, the amount of infectious virus and viral RNA copies that were recovered behind the facemask were respectively on average 2.3- and 2.4-logs lower than in the no-mask control in the first session. This corresponds to filtration efficiencies of 99.4 and 99.6% (**Fig. 4a and b**). When this type of facemask was used in the second session, the infectious virus and viral RNA recovery decreased by respectively 3.1- and 3.5-logs on average, which corresponds to 99.92 and 99.96% filtration efficiency (**Fig. 5a and b**). The Reinier 0.1-facemasks reduced the recovery of infectious virus and RNA copies on average by 2.1 and 2.8-logs respectively compared to the no mask-control. This corresponds with the filtration efficiencies of 99.2 and 99.9% (**Fig. 4**). 2.3 and 2.6-log reductions were observed with the DSM 1.0-facemasks, which corresponds with 99.3 and 99.6% filtration efficiency. The Reinier 1.0-facemasks were tested in the second session, which reduced the infectious virus and total viral RNA copy numbers with 3.1- and 2.6 logs on average, corresponding to 99.92 and 99.71% filter efficiency (**Fig. 5**).

**Fig 4.**
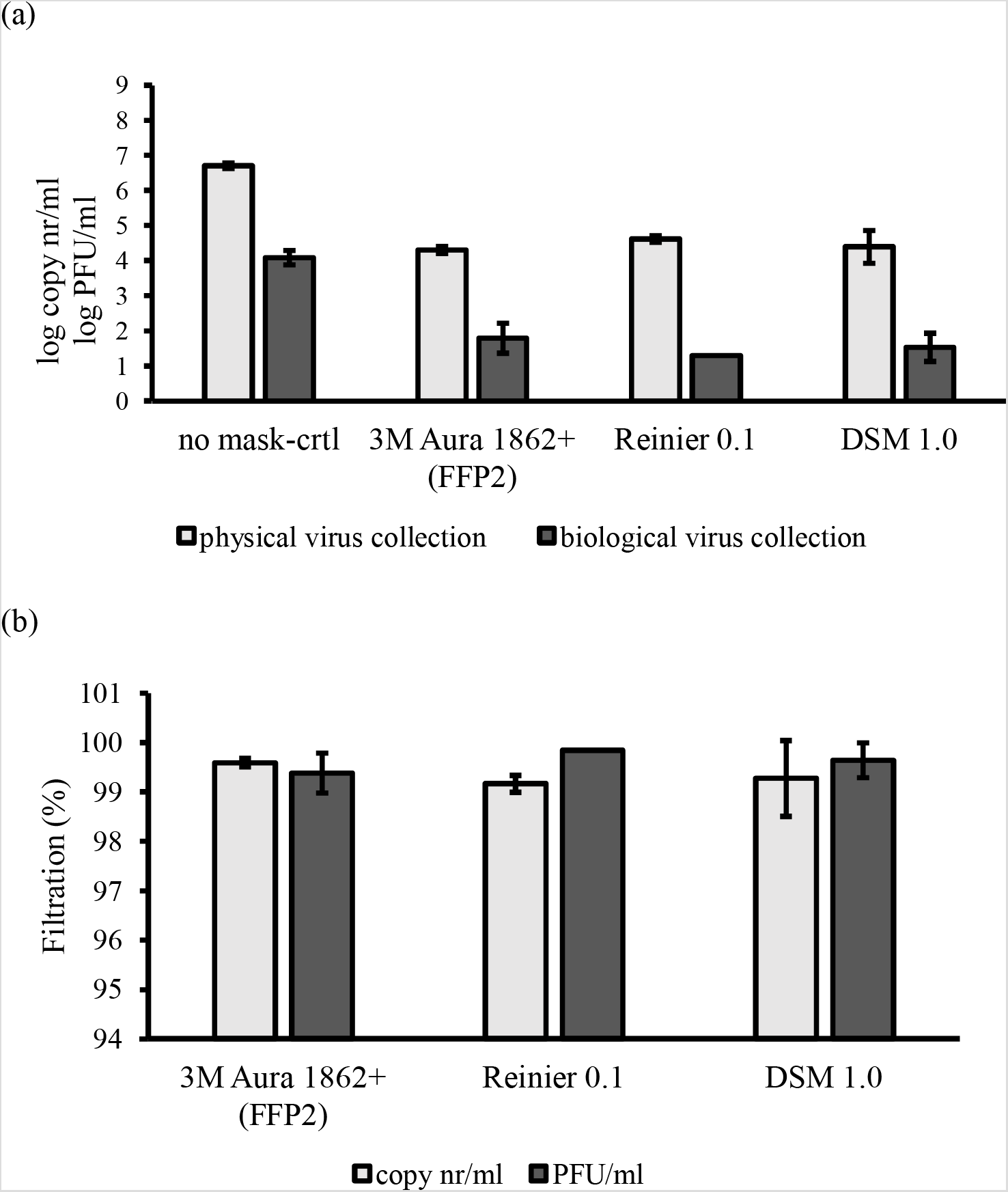
Physical and biological collection of aerosolized mouse hepatitis virus (a) and filtration efficiencies (b) observed for facemask Reinier 0.1 or DSM 1.0.

**Fig 5.**
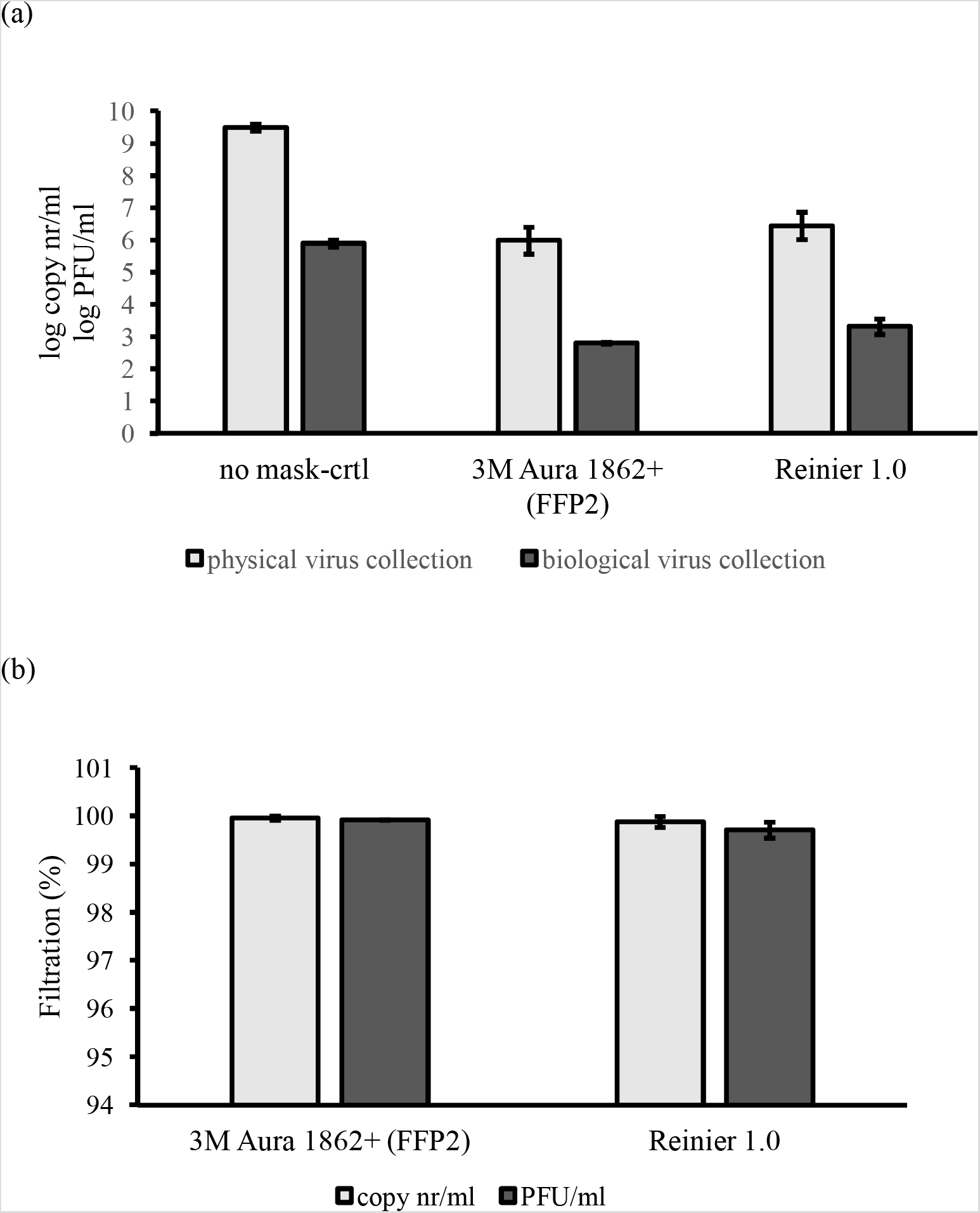
Physical and biological collection of aerosolized mouse hepatitis virus (a) and filtration efficiencies (b) observed for facemask Reinier 1.0

The virus filtration efficiency of the locally-produced facemasks was determined at a continuous air velocity of 0.42 m sec^−1^, which is significantly higher than during the above described NaCl particle penetration test. Similar particle filtration efficiencies were nevertheless observed in these two experiments. This air velocity is also higher than described in various other studies where the virus filtration efficiencies of facemasks were tested (Borkow et al., 2010, Harnish et al., 2013, 2016, Rengasamy et al., 2017, Zhou et al., 2018). It was chosen here to test at 0.42 m sec^−1^ as this is a more physiological relevant air velocity during inhalation as the maximal air velocity during an inhalation cycle reaches up to 1 m/sec (Tang et al., 2013). Although bacteriophages, influenza viruses, and rhinoviruses have previously been used in filtration experiments (Borkow et al., 2010, Harnish et al., 2013, 2016, Rengasamy et al., 2017, Zhou et al., 2018), here a coronavirus was used for the first time. Despite the higher air velocity and the usage of a different virus, at least 98% of the virus-loaded aerosols were filtered by the FFP2-certified facemask, similarly as observed in the prementioned studies. The locally-produced facemasks showed similar virus filtration efficiencies as the FFP2-certified facemask, which suggests that these are equally protective against coronavirus-loaded aerosols.

### Reinier-0.1 and –1.0 facemasks have acceptable inward leakage around the edges

In addition to the filtering capacity of the facemask’s filter material, a proper fit is also crucial for the protective quality of the facemask (Rengasamy et al., 2011, Serfoze et al., 2017). It had been modeled by others that when the area of inward leakage at the facemask’s edges exceeds 0.1% of its total surface area, the facemask is unable to offer 95% protection (Mukhametzanov et al., 2016). Therefore the three locally-produced facemasks were fit-tested to estimate their protective quality when used by individuals. Those individuals performed various exercises while wearing the facemask as described in material and methods. FFP2-certified facemasks are allowed to have an average inward leakage of maximal 8 or 11%, depending on the number of tests and exercises (**Table 1**). As our facemasks were tested by only three different persons, not the average, but maximal observed inwards leakage was taken as a measure for the fit of a facemask (**Table 2**), thus defining a worst-case scenario. The FFP2-certified facemasks showed a maximal inward leakage of 0.5% (**Table 2**). The Reinier 1.0-facemasks showed the best fit of the self-designed masks with a maximal observed inward leakage of 0.8%, whereas the DSM 1.0-facemasks showed a maximal inward leakage of 14.6%. The Reinier 0.1-facemasks showed a maximal inward leakage of 4.8%. These observations indicate that based on three study subjects, the Reinier-0.1 and −1.0 models have acceptable fits.

**Table 2:**
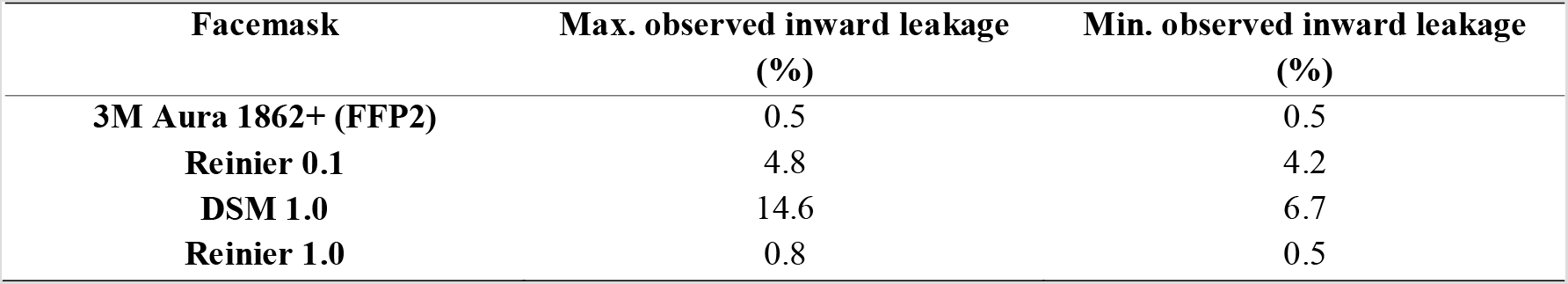
Maximal and minimal observed inward leakage

| **Facemask** | **Max. observed inward leakage** | **Min. observed inward leakage** |
| --- | --- | --- |
|  | **(%)** | **(%)** |
| **3M Aura 1862+ (FFP2)** | 0.5 | 0.5 |
| **Reinier 0.1** | 4.8 | 4.2 |
| **DSM 1.0** | 14.6 | 6.7 |
| **Reinier 1.0** | 0.8 | 0.5 |

### The locally-produced facemasks can be safely used, regarding breathing resistance

The breathing resistance of the facemasks was determined to obtain an indication about the wearer’s comfort and safety when wearing these facemasks. Therefore, the facemasks were placed on a manikin head and the pressure drop was recorded while the airflow was increased. The maximal pressure drop at an inhalation flow of 30 or 95 L min^−1^ according to the NEN-149 standards is 0.7 and 2.4 mbar respectively. The pressure drops varied between 0.14 and 0.16 mbar at a continuous flow of 30 L min^−1^, and at a flow of 95 L min^−1^, the pressure drop varied between 0.74 and 0.85 mbar for all three facemask models (**Table 3**). Exhalation at a continuous flow of 160 L min^−1^ resulted in a pressure drop of between 1.11 and 1.44 mbar for the three different designs, which is below the 3.0 mbar that is allowed according to the NEN-149 standards. These observations indicate that all three designs meet the NEN149 standards regarding breathing resistance. However, for the DSM-facemask this result might be due to the significant amount of inward leakage.

**Table 3:** Pressure drop over the self-designed facemasks at continuous inhalation flows of 30 or 95 L min^−1^ and an exhalation flow of 160 L min^−1^.

| Facemask | Inhalation<br>30 L min<sup>-1</sup><br>(0.7 mbar max.) | Inhalation<br>95 L min<sup>-1</sup><br>(2.4 mbar max.) | Exhalation<br>160 L min<sup>-1</sup><br>(3.0 mbar max.) |
| --- | --- | --- | --- |
| Reinier 0.1 | 0.14 ± 0.003 | 0.74 ± 0.009 | 1.44 ± 0.05 |
| DSM 1.0 | 0.14 ± 0.003 | 0.84 ± 0.008 | 1.31 ± 0.06 |
| Reinier 1.0 | 0.16 ± 0.002 | 0.85 ± 0.01 | 1.11 ± 0.06 |

## CONCLUSION

It is concluded that the Reinier 0.1 and –1.0 facemasks provide good respiratory protection against coronavirus-loaded aerosols and with similar efficiency as a certified FFP2 mask, suggesting that these can be safely used by healthcare professionals.

## Data Availability

All obtained data are in the manuscript file and supplemental data file

## ACKNOWLEDGMENTS

This work was supported by the NIH/NIAID (contract number HHSN272201400008C) and European Union’s Horizon 2020 research and innovation program VetBioNet (grant agreement No 731014). Part of this research was supported by the Leiden University Fund (LUF), the Bontius Foundation, and donations from the crowdfunding initiative “wake up to corona”. S.H. was funded in part by an NWO VIDI grant (contact number 91715372). Part of this research was supported by ZonMw initiative “creative solutions against CoVID-19” (5001-0013). The authors also wish to thank Monique Elsing-van Olphen for supporting this research and for supplying information on the facemask designs.

